# Estimating the gendered impact of COVID-19

**DOI:** 10.1101/2022.11.02.22276012

**Authors:** Samer Singh, Rakesh Kumar Singh

## Abstract

The extent of gendered COVID-19 impact remains undetermined for the lack of sex-disaggregated data. The prevailing view puts males nearly twice as impacted as females. Globally, access to resources and their usage are gendered- mostly favoring males. Gender gaps widen during natural/man-made calamities and pandemics. Modeling estimates of impact for top 70 countries reporting >300 sex-disaggregated COVID-19 deaths (>80% of total), indicates average mortality sex (male:female) ratio (COVID-MSR) of 1.37±0.30 (95% confidence interval:1.30-1.44; range:0.85-2.47) against prevalent pre-pandemic MSR of 1.79±0.41 (1.70-1.89; range:0.93-2.99). Contrary to the prevailing view, widened gender gaps globally increased female mortality by 19.57±21.16% (14.62%-24.88%; range: -22.46 to +68.50%) causing an estimated 22.03% excess deaths (360 thousand by 30 December 2021). Identification of factors favoring gendered impacts is needed for equitable pandemic management.

**One-Sentence Summary:** **‘Missing Females’ in COVID-19**?

## INTRODUCTION

As of 30 December 2021, 288.95 million COVID-19 cases and 5.47 million associated deaths had been reported *(1)*. The cases and deaths had been more in middle- and high-income countries as compared to low-income countries. Generally, up to two-fold higher COVID-19 mortality for males had been assumed based on early-stage pandemic data from small-scale studies *(2,3,4)*. The supposed inequity of COVID-19 mortality and disproportionate deaths of males has been concerning. The attribution of high male mortality to various biological differences *(5)* would have promoted the undertaking of research efforts and policies guided by such assumptions, largely overlooking the general global prevalence of high male mortality in adults as compared to females (up to 3-fold, global average 1.5-fold) stemming from underlying socio-structural factors, and risk-taking behaviors *(6)* despite more control exercised by males over the resources and preferential treatment *(5,7)*. A comparative detailed country-specific analysis of gendered COVID-19 deaths with that of pre-pandemic levels of gendered mortality for the estimation of the magnitude of gendered impact is needed to accurately guide the management practices for an equitable balanced response - mitigating the effect of underlying potential risk factors adversely affecting one gender over other. Emergencies and pandemics are known to change the mortality sex ratios (MSR) and their estimates can be used as a guide to identifying underlying structural deficiencies and response gaps to attain equity *(6)*. However, the general lack of global sex-disaggregated data since the beginning of the COVID-19 pandemic had prevented any large-scale estimation of country-wise equity gaps for comprehensively guiding the local policies toward achieving a balanced response.

## RESULTS AND DISCUSSION

The data collated by the site Global Health 50/50 from different governments as and when available had been a single source of COVID-19 related sex-disaggregated information *(8)*. The release of sex-disaggregated data had been generally delayed (2-10 months) and only accounted for a fraction of the total COVID-19 mortality barring a few developed countries of North America and Europe (**Table S1** as of 15 September 2021). For the top 70 countries reporting at least >300 sex-disaggregated data, the estimated COVID-19 cases sex (Male: Female) ratio (CSR) ranged from 0.67 (Ukraine) to 2.45 (Bangladesh). Surprisingly, more female cases (CSR<1) were reported in Europe and the Americas while male cases (CSR>1) were in Asian, African, and American countries near the tropics (**Fig. 1A**). The reported COVID-19 mortality rate sex ratio for all cases (COVID-19 MSR) had been generally higher than unity (**Fig. 1B, Table S1**). Males appeared to have experienced more COVID-19 mortality, with only 5 countries out of the top 70 countries reporting sex-disaggregated data had their MSRs below one (Australia 0.95, South Africa 0.95, Moldova 0.96, Slovenia: 0.96, South Korea 0.99; colored purple in **Fig. 1B**). The highest MSR of 3.35 was recorded for Bangladesh and the lowest of 0.95 for Australia and South Africa, while the global MSR average for countries (included in the analysis) was 1.48±0.47 (95% Confidence interval UL: 1.59; LL: 1.37; **Table S1**).

**Fig. 1.**
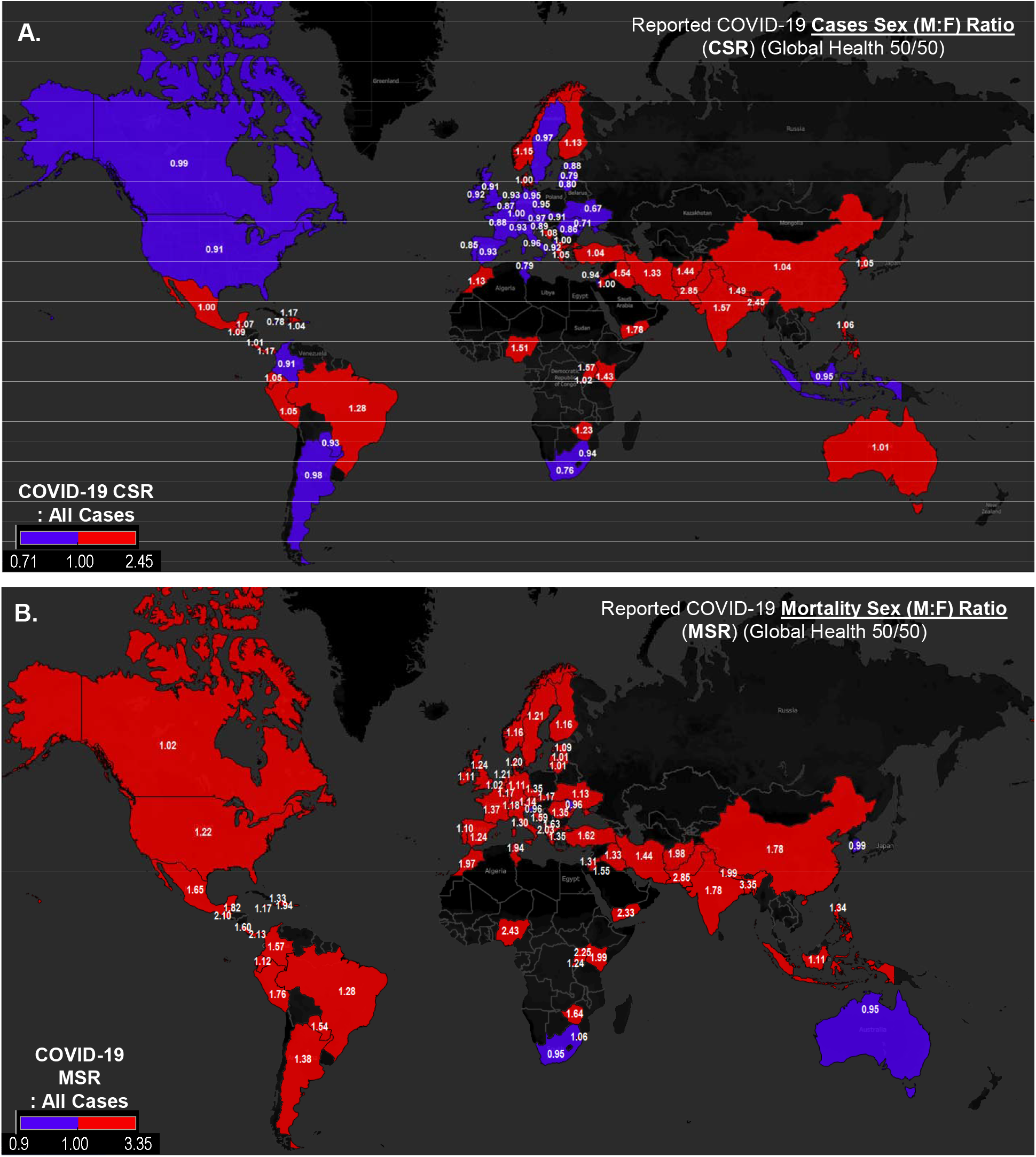
Reported COVID-19 Cases and Mortality display gendered impact. **(A)** Reported COVID-19 cases sex ratio (CSR) for countries was more than 1 (red color) in mostly middle- and lower-income countries of Asia, Africa, and the Americas near the equator while less than 1 (purple) in High-income countries of Europe, North America and some countries in South America, Africa, and Southeast Asia. **(B)** The reported COVID-19 mortality sex ratio (MSR) for most countries is more than 1 (red color) with a few exceptions in Europe Asia Africa and Australia (purple color) indicating disproportionately higher male deaths in most countries (See **Table S1**).

The reporting of COVID-19 positive cases has been differentially affected by changing guidelines, diagnostic methods, and kits and their cutoffs adopted/employed during the pandemic *(9,10)*. The World Health Organization (WHO) recommended RT-qPCR based COVID-19 diagnostics are considered ‘gold standard’ for their higher sensitivity and specificity *(11,12)*, whereas other methods were employed/approved for use to fill the supply gap and decrease the turnaround time to manage the pandemic despite them having lesser sensitivity and specificity-many a times inflating false positives as high as twice under various circumstances (*9-10,13*). To arrive at more reliable estimates of gendered COVID-19 impact on populations and reduce the imprecision in ascertaining the gap in gender equity, the analysis of only RT-qPCR confirmed cases was undertaken.

The COVID-19’s MSR estimate based on RT-qPCR positive cases only (*i*.*e*., COVID-MSR) indicates a disproportionate gendered impact - male mortality lower than unity in only 4 countries out of the top seventy countries reporting sex-disaggregated deaths while 66 countries reported more male deaths. India leads the minority pack of countries, with an MSR of 0.85, displaying higher female deaths as compared to males in RT-qPCR confirmed cases ‘RPCC’ (**Fig. 2A; Table S2**). Other countries closely following India in more female deaths (MSR<1.00) were Iraq (0.87), South Korea, and Australia (0.95). The group of countries with higher male deaths was led by Tunisia with the highest MSR estimate of 2.47, while global COVID-MSR for all countries averaged 1.37±0.30 (UL: 1.44; LL: 1.30; **Table S2**). These values suggest COVID-19 had been more averse to males.

**Fig. 2.**
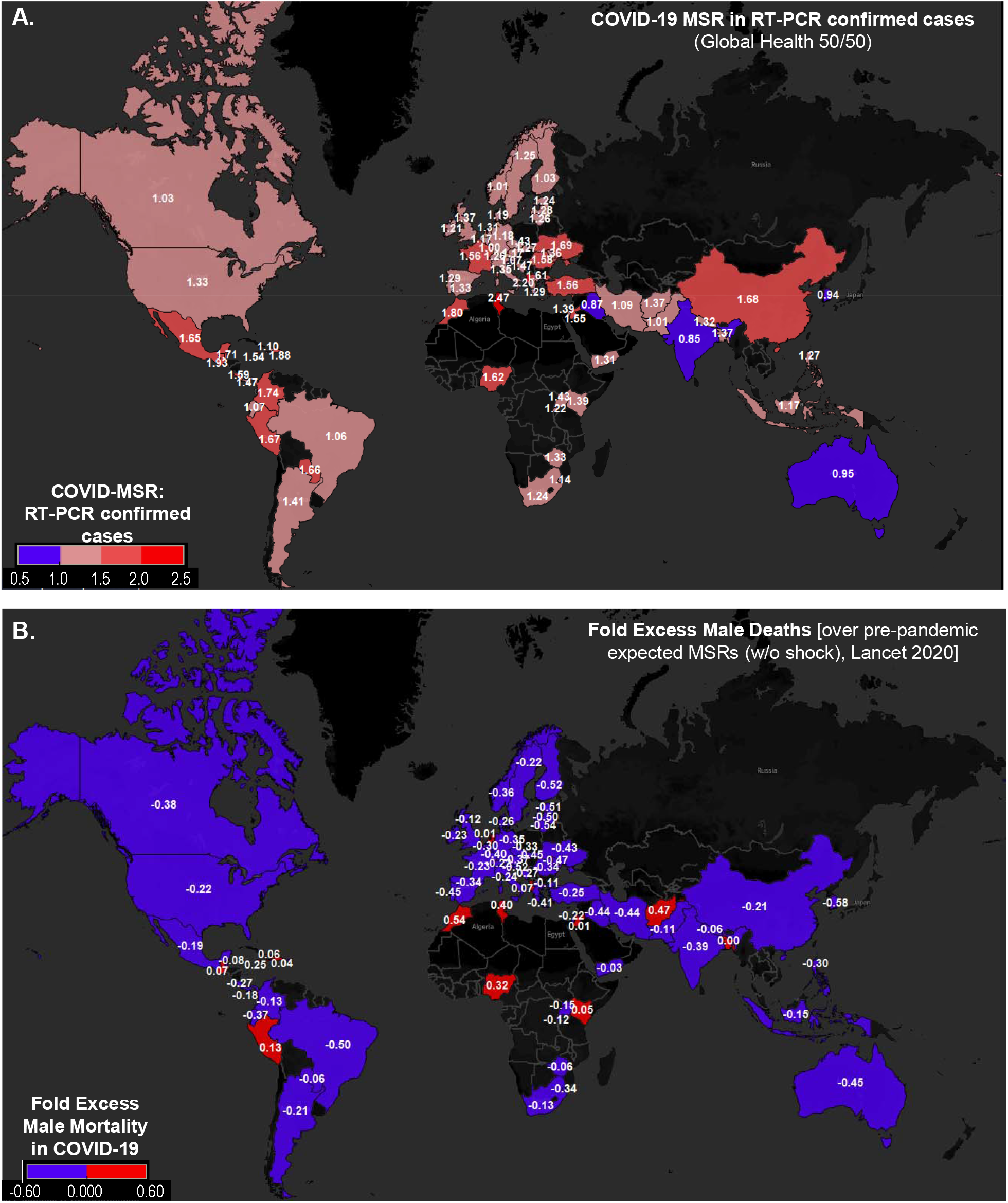
Overwhelmingly COVID-19 Mortality Sex Ratio (MSR) in RT-PCR confirmed COVID-19 cases (RPCC) were lower than pre-pandemic MSR of countries. **(A)** The MSR in RPCC for countries was lower than all methods diagnosed cases (see **Fig. 1**). Only India, Iraq, Australia, and South Korea had higher female mortality rates (colored purple) while others had higher male mortality rates (colored red, associated MSR in white). **(B)** Most countries had negative fold excess male mortality, i.e., [{(COVID-MSR)/(MSR w/o shock)}-1]. The estimated COVID-MSRs in RPCC have been lower than the pre-pandemic estimate of MSR ‘w/o shocks’. The countries were colored purple and red for negative and positive deviation, respectively (associated fold excess male mortality indicated in white). Except for a few countries of Asia, Africa, and South and Central America, (colored red) from the lower-income group, most countries had lower COVID-MSR (purple) or lesser male mortality rates than background levels. Note, Bangladesh which reported the most gendered COVID-19 impact, reporting 3.35-fold higher male mortality in COVID-19 came out to have the least gendered impact (almost zero-fold excess male mortality or deviation from anticipated MSR (See **Table S2**).

Globally, substantial sex differences in the adult mortality rates or MSRs exist among the countries influenced by education, sociocultural factors, demographics, developmental index, etc. *(6,14,15)* The country-/region-specific estimates are driven by variables unique to them. Usually, their variations during adverse periods including pandemics reflect the changes in the underlying contributory factors that may be favoring one gender over the other *(6)*. Their variation is a good indicator of changing dynamics and points to the requirement of balance restoration. The elderly and aged had been most severely impacted (>10,000) by COVID-19 as compared to <15-yr.-olds. The MSR usually varies with age and overall shows an increasing trend with age. It reaches a value between 2-3 for adults (15-60 yr.) for many countries included in the analysis [See **Table S2** and *(6,16)*]. Considering the paucity of age-segregated COVID-19 mortality data for all countries, differences in demographic composition and the median age along with the most affected happened to be the comorbid adults or elderly/aged, the adult MSRs could be considered a fair approximation for estimating the gendered impact of COVID-19 (though could be potentially underestimating the issue!). The latest sex-specific adult mortality rate estimates from the Global Health Observatory (GHO, WHO, 2018) *(14)* and a recently updated estimates from the Global Burden of Diseases, Injuries, and Risk Factors Study (GBD) 2019, that show similar differential mortality rates specific to countries with some countries-specific adjustments for the change in the underlying influencing factors *(6, 16)* (**Table S2**) are available for immediate pre-pandemic times. As per estimates for the year 2019, globally, on an average 50% higher mortality was observed among adult males as compared to females *(6,16)*. Countries of regions, Europe & North America, and Latin America & Caribbean had the worst extreme average MSRs of 2.1 and 1.9, respectively *(6,16)*. The countries of Sub-Saharan Africa with the younger population, lower median age, and life expectancy, displayed the lowest MSR of 1.2, *i*.*e*., just 20% more mortality rate among males as compared to females *(6,14)*. The adjusted adult mortality rates or corresponding MSR with corrections for man-made disasters/discontinuities for the countries (e.g., war, famine, terrorism), or ‘MSR w/o shocks’ *(6)* would be more appropriate for estimating the gendered impact of COVID-19 on the global population. The MSR values for 2019 (w/o shocks) could be used as a benchmark for ascertaining the gendered impact on populations.

The average pre-pandemic ‘MSR w/o shocks’ for the 70 countries included in the current analysis was 1.79±0.41 (UL: 1.89; LL: 1.70; range: 0.93–2.99; **Table S2**). Surprisingly, the COVID-19 specific MSR (COVID-MSR) estimate for the same countries based on confirmed RT-qPCR positivity [average 1.37±0.30 (UL: 1.44; LL: 1.30)] have been overwhelmingly lower for most of them (see **Table S2** with **Table S1**). The estimates of average relative change in MSR for COVID-19 as compared to expected MSR w/o shock estimate for the year 2019 for the group of countries investigated in the current report (i.e., reporting at least more than 300 sex-disaggregated deaths) come negative with the value of -0.2±0.24 [UL: -0.14; LL: -0.26; range: - 0.58 (South Korea) to +0.54 (Morocco); **Table S2**]. The deviation of average COVID-MSR estimate for the countries came close, *i*.*e*., -0.18±0.26 (UL: -0.12; LL: -0.24; **Table S2**) even when the adult MSR estimate from WHO, 2018 *(14)* was used for the comparison. A quick glimpse of COVID-MSR for countries (**Fig. 2A**) and its relative deviation from the expected pre-pandemic adult MSR for countries (**Table S2**) indicates overwhelmingly disproportionate excess death of females rather than males. The estimated fold excess male mortality for countries indicates overwhelmingly lower male mortality in the majority of nations (colored purple in **Fig. 2B**).

As MSR for countries varies widely, just looking at COVID-MSR deviation from the anticipated or fold excess mortality could not accurately portray the magnitude of gendered impact on the population. So, the excess male and female COVID-19 deaths for countries were calculated over that anticipated from countries’ pre-pandemic MSRs to calculate the gravity of the situation (Supplementary Material & Methods; **Table S3**). The estimation of excess gendered COVID-19 deaths for countries that would be expected from pre-pandemic MSR estimates indicated on an average about 10% lesser deaths in males [-9.38%±10.70%; UL: -6.88%, LL: -11.89%; range: - 29.99% (South Korea) to +19.96% (Afghanistan); **Table S3**]. Whereas average excess deaths observed in females was about 20 % [19.57%±21.16%; UL: +24.88%, LL: +14.62%; range: - 22.46% (Morocco) to +68.50% (South Korea)]. The variation of % excess male deaths with that of the females for the countries is presented in **Fig. 3A**. Overall, only fourteen countries out of a total of seventy (14/70), mostly from the tropics with lower human developmental index (HDI) and median age (**Table S4**) have observed excess male deaths while others have experienced more female deaths. The higher % male deaths reported in some of the lower HDI countries could also result from a general higher propensity of the occurrence of medically uncertified deaths among women than men in poorer places as also recently reaffirmed by a COVID-19 study in India (*15*). So, the actual COVID-MSR increase over the existing estimate of w/o shock MSR for these countries could be still an overestimate of the male-gendered impact. These estimates could get corrected in the future with the release of complete sex-disaggregated mortality data by countries.

**Fig. 3.**
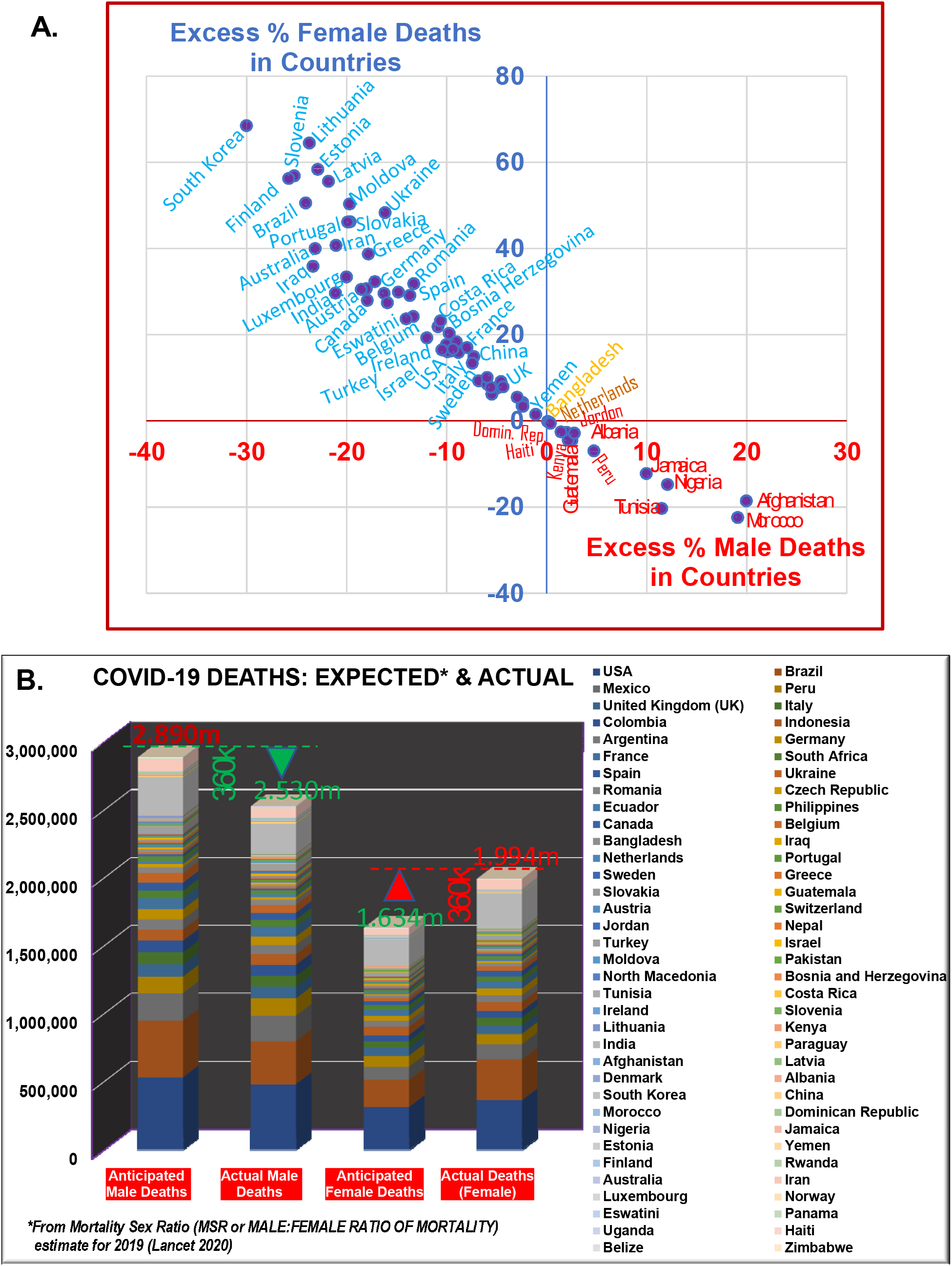
Estimates of gendered COVID-19 impact on countries. **(A)** Relative % excess gendered COVID-19 mortality in different countries than that anticipated from pre-pandemic MSRs of countries. Exclusive higher % excess female mortality was observed in high-income countries (<14,601 USD GDP per capita; **Fig. S1**), whereas the % excess male deaths were confined to lower-income countries. **(B)** Graphical representation of estimates of anticipated sex-specific mortality *vs* actual COVID-19 deaths for countries (as of 30 December 2021). Overall higher female mortality rates (average 19.57% ±21.16%; UL: +24.88%, LL: +14.62%) among countries caused an estimated 360 thousand excesses, i.e., 22.03% more female deaths while 12.46% lesser male deaths, as would have been anticipated from prevalent pre-pandemic adult MSR for countries (**Table S3**).

**Fig. 3.**
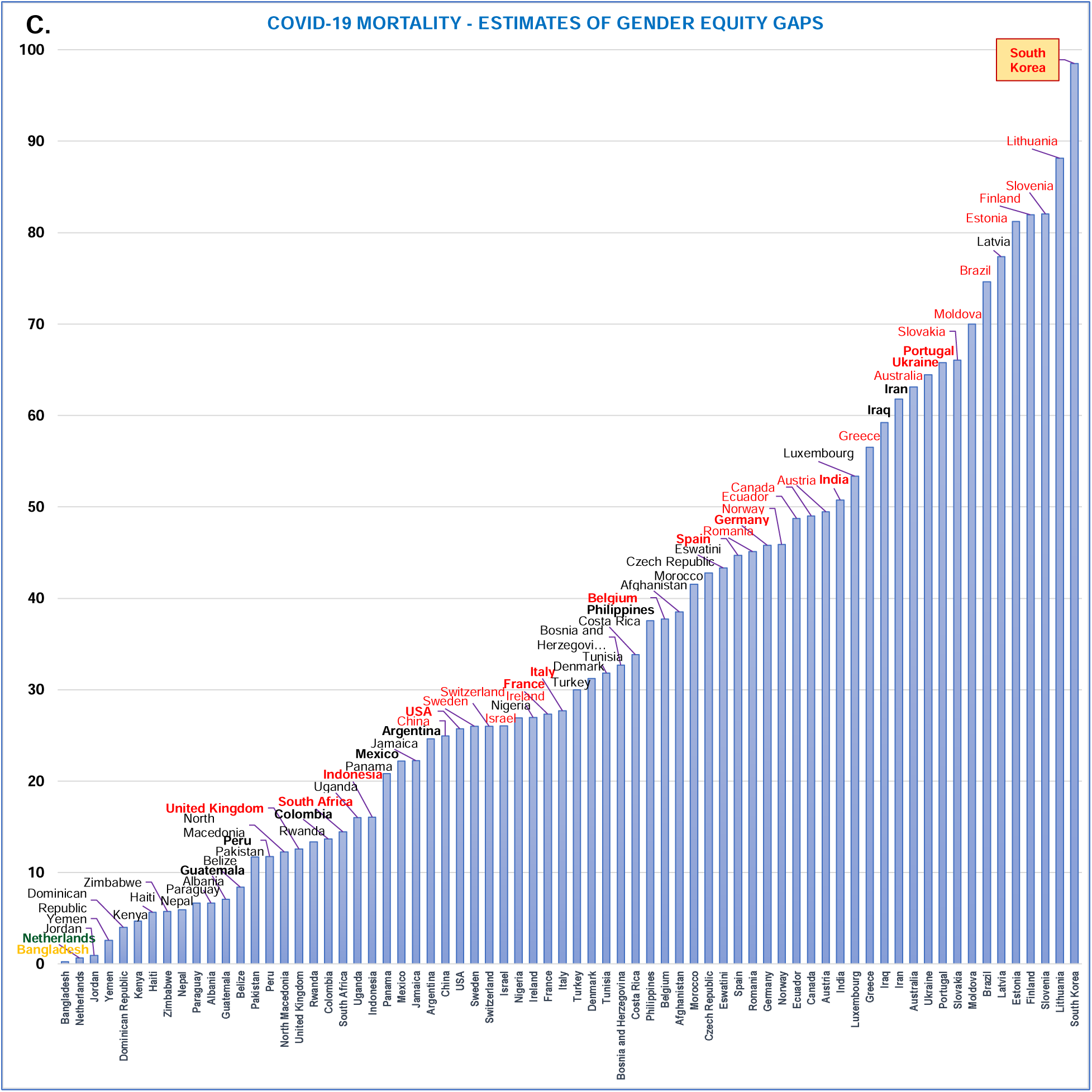
**(C)** The observed relative gender equity gaps potentially reflect the gendered response of countries during COVID-19. ***Note:*** Bangladesh, the Netherlands, and Jordan seemed to have the most gender-neutral response. Others whether from low- or high-income group countries seemed to have varying levels of gendered response - most of the time putting females in a disadvantageous position (see **Table 1**).

The extrapolation of country-wise gendered COVID-19 death estimates to COVID-19 data on 30 December 2021 for these countries indicates the gendered response would have resulted in 22.03% more female deaths while 12.46 % lesser male deaths in these countries (**Table 1**). Overall, estimated 360 thousand excess female deaths or ‘missing women’ during the COVID-19 pandemic over and above 1634 thousand deaths anticipated/expected from the country-specific MSRs had ensued in these countries (**Table 1**). Male deaths were similarly 360 thousand lesser than the anticipated 2890 thousand had the country-specific MSRs been maintained/observed for COVID-19. The anticipated and actual (projected) COVID-19 gendered deaths for these countries are graphically presented in **Fig. 3B**.

**Table 1.**
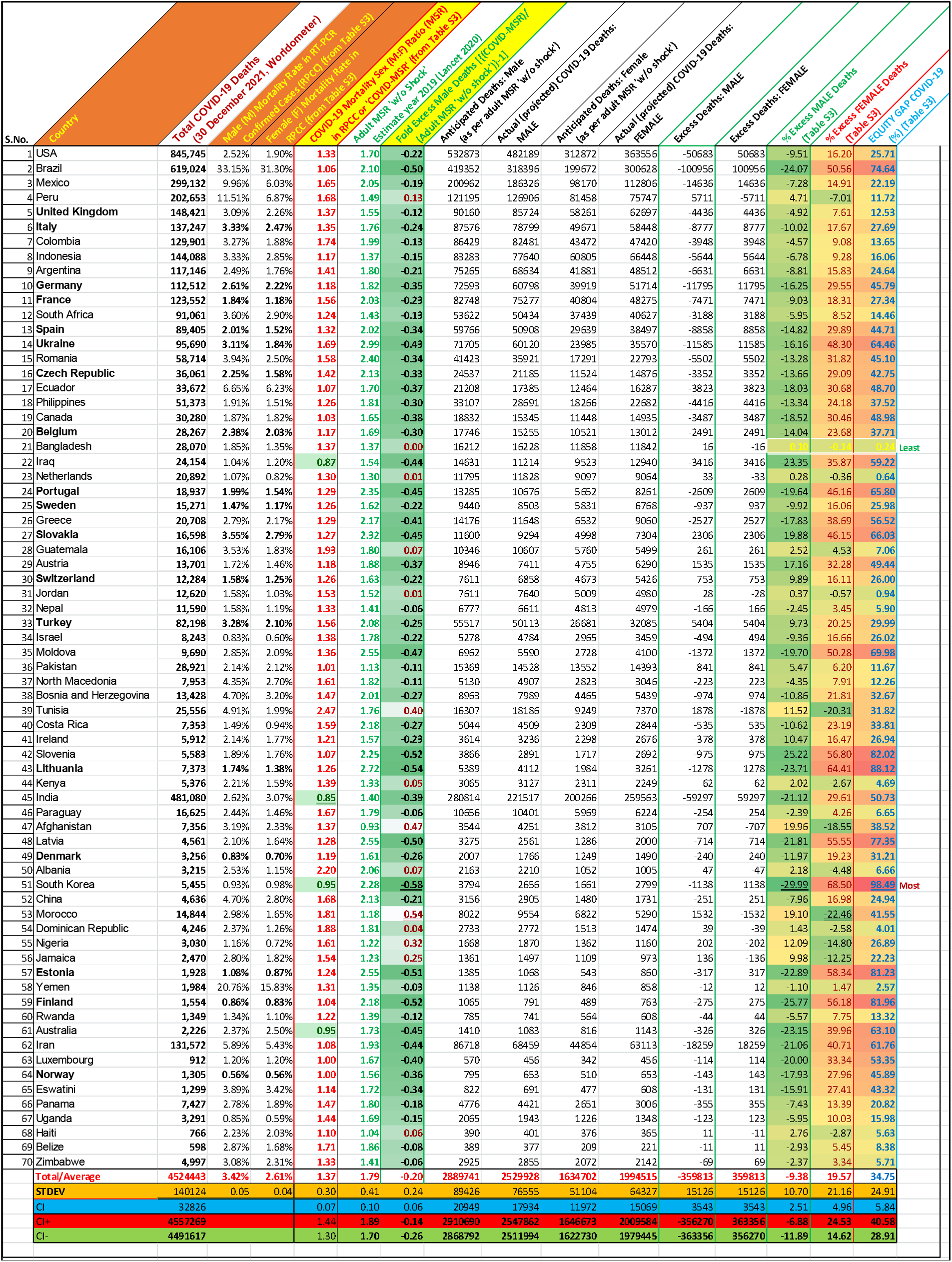
Estimate of “Missing Women”, and Gender Equity Gaps in COVID-19: Modeled estimates of gendered impact on countries [Excess Deaths: COVID-19 specific gendered mortality - anticipated and actual (projected)] as of 30^th^ December 2021 and observed gender gaps. The relative gendered response estimates are based on the change in sex-specific mortality during COVID-19 (COVID-MSR) as compared to country-specific pre-pandemic MSRs (**Table S3**). (CI: 95% Confidence interval; CI+: Upper limit; CI-: Lower limit). For calculations see Supplementary Methods.

Any sudden change in MSR during epidemics/pandemics usually represents response variation for populations during an event and is generally an indicator for the direction of course correction potentially required to again return to supposed normalcy (equal to response in conditions without shocks) from the stressed response. The existence of wide-ranged deviations of the COVID-MSR, both positive and mostly negative, from the populations’ pre-pandemic MSR w/o shocks (**Table 1, Table S3**), potentially indicates a general disproportionate gendered response to the medical needs and required support among countries not necessarily arising from biology favoring or disfavoring one sex over the other *(5)*. Based on different MSR rates for countries, the changed mortality rate for genders (or MSR) translates into a differential equity gap for countries. The overall equity gap, the summed-up deviation of gendered excess deaths as per COVID-MSR estimates (**Table 1**, last column), is presented graphically in **Fig. 3C**. Surprisingly, Bangladesh which reportedly happened to have displayed the most gendered impact as per Global Health 5050 reported COVID-19’s MSR of 3.35 (28k deaths), along with the Netherlands (MSR: 1.21, 21k deaths) appeared to be the most gender-neutral countries in COVID-19 response. Bangladesh, Netherlands, and Jordan displayed the least COVID-19 specific equity gap (<1 point), potentially indicative of a minimal change in their response during COVID-19. Except for Jordan and Netherlands, all countries showing less than a 10-point variation in the equity gap are from the lower HDI and GDP per capita countries (**Table S4, Fig S1 & S2**). Better-off countries had displayed larger COVID-19 equity gaps, with an equity gap of 98.49 points South Korea topped the list among the countries investigated. Among the top 11 countries reporting COVID-19 cases of total *(1)*, the associated equity gap was as follows (% cases % of total global; estimated equity gap points): USA (16.20%; 25.71), India (8.41%; 50.73), Brazil (5.94%; 74.64), France (5.58%; 27.34), Germany (4.82%; 45.79), United Kingdom (4.30%; 12.57), Russia (3.54%; ND), South Korea (3.35%; 98.49), Italy (3.20%; 27.69), Turkey: 2.93%; 29.99), Spain (2.32%; 44.71). Though Brazil, India, and the USA displayed gender gaps about three-quarter, two-quarters, and one-quarter of South Korea, these countries due to large number of total deaths (*i*.*e*, Brazil: 619 thousand (k), India: 481k, USA: 845k), are estimated to have been home to more than half of the ‘missing women’ globally from COVID-19, accounting for approximately 101, 59 and 51 thousand excess female deaths, respectively. The extreme equity gaps displaying countries (in decreasing order) had been generally smaller with lower total cases and associated mortality, *e. g*., South Korea (5k deaths), Lithuania (7k deaths), Slovenia (6k deaths), Finland (2k deaths), Estonia (2k deaths).

Our estimates of gendered COVID-19 impact on countries could still be a gross underestimate stemming from non-equitable resource allocation, and quality control differential in diagnostic and reporting biases across countries. The extent of diagnostics’ quality and reporting bias is potentially reflected by the differential in reported gross COVID-19 MSR (all methods) *vs* RT-PCR confirmed COVID-19 cases or COVID-MSR (1.48 *vs* 1.37; **Table S1 & S2)**. Surprisingly the fold differential is generally insignificant to small in the range of 1 to 1.25 fold for developed high- and middle-income countries, e.g., USA (all method MSR *vs* RT-PCR confirmed MSR; 1.22 *vs* 1.33), UK (1.24 *vs* 1.37), Germany (1.11 *vs* 1.18), France (1.37 *vs* 1.56), South Korea (0.99 *vs* 0.95), Jordan (1.55 *vs* 1.53), Morocco (1.97 *vs* 1.81), Albania (2.03 *vs* 2.20), Tunisia (1.94 *vs* 2.47), but generally large for lower-income countries displaying the difference in the range of 1.2 to >2.5 fold, e.g., Brazil (1.28 *vs* 1.06), **India (1.78** *vs* **0.85), Pakistan (2.85** *vs*, **Bangladesh (3.35** *vs* **1.37)**, Afghanistan (1.98 *vs* 1.37), Guatemala (2.10 *vs* 1.93), Nigeria (2.43 *vs* 1.61) Dominican Republic (1.94 *vs* 1.88), Peru (1.76 *vs* 1.68), Kenya (1.99 *vs* 1.39), Haiti (1.33 *vs* 1.10). *Surprisingly, the COVID-MSRs increased for countries of Europe and North America when only RPCC was considered while it decreased for other countries in comparison to estimates of COVID-MSR from all methods. Whether it only reflects reporting disparities arising from differences in policy and the access to quality diagnostics or gender differences in health-seeking behavior and preferential treatments in the different socioeconomic contexts of countries, is something that the countries would like to investigate for course-correcting their disaster management practices and preparedness*. Furthermore, the observed higher male mortality in lower-income group countries could be an artifact of higher stress borrowing common for males and the prevalence of more frequent quality medical access and treatment preference, whereas many female deaths remain uncategorized in poor settings*(17)*. The country-specific factors driving the gendered impact need to be identified for the implementation of proactive steps to allow more equitable resource allocation, distribution, access, and usage to minimize the widening of the gender gap during stressed situations. Previous studies have modeled the contribution of different variables on COVID-19 impact on populations for identifying explanatory as well as protective variables *(18-21)*. An exploratory covariation analysis and modeling of the disproportionate gendered death (male/ female) for countries with such variables could be an interesting area of investigation to identify potential variables responsible for gendered impacts (**Fig. S1 & Fig. S2**).

## CONCLUSION

Overall, our study indicates females have been disproportionately affected by the COVID-19 pandemic, registering about 22% excess mortality or 360 thousand ‘missing females’ in the analyzed top 70 countries (accounting for >80% of COVID-19 deaths), contrary to the prevailing notion of males being disproportionately affected. It highlights the existence of potential COVID-19 specific gender gaps disfavoring females. Further internal audits and reassessment of sex-disaggregated data in countries wherever available are warranted to corroborate and validate the extent of gendered COVID-19 impact. It may require an upward revision of the number of missing females in the COVID-19 pandemic. The analysis of sex-disaggregated quality data with potential contributors could help identify the country-specific variables leading to skewed gendered impact. Identification of potential contributory factors could allow readjustment of priorities to fill the gender gap created by COVID-19. The policies guiding COVID-19 management may have to be consciously shifted to attain gender neutrality to minimize the gendered COVID-19 impact. The steps taken now could help countries be better equipped for any such eventuality in the future to respond equitably and allow for helpful resource distribution to minimize gendered impact.

## MATERIALS & METHODS

### MATERIALS

#### Data Collection

##### Sex-specific COVID-19 incidence and mortality data

The country-specific sex-disaggregated COVID-19 incidence and mortality data collated from specific countries’ government and health websites by globalhealth5050.org *(8) was* collected/accessed on 15 September 2021 is shown in the **Table S1**. For United Kingdom (UK) the data available for England, Wales, Scotland and Northern Ireland was summed up as and used as such representing UK (the total case and mortality counts reported for UK as whole at Worldometer *(1)* for indicated date were little less than the sum of the parts reported, possibly resulting from duplication of counts in different parts or data reporting/collation differences by different agencies.

As RT-PCR based diagnostics is considered ‘gold standard’ for SARS-CoV-2 detection *(11,12)* for its superior sensitivity and specificity, the mortality in RT-PCR positive gender identified data set reporting at least more than 300 sex disaggregated deaths for statistical considerations were included in current analysis (**Table 1**).

##### Pre-pandemic and COVID-19 specific Adult Mortality Sex Ratio

The sex-specific adult mortality is an important indicator for underlying factors overall impact on the mortality pattern of a population *(14)*. During stressful conditions, they mostly change as a result of widening of gender gaps - disfavoring females. Adult mortality is defined as “probability that a 15 year old die before reaching his/her 60^th^ birthday (per 1000 population)… per year among a hypothetical cohort of 100,000 people that would experience the age-specific mortality rate of the reporting year” *(14)*. The recent sex-specific mortality rates estimate for adults (15 -60 years) were obtained from following: (i) WHO 2018 (Global Health Observatory) latest available for year 2016 *(14)* and (ii) GBD demographics collaborators 2019 published in Lancet 2020 for year 2019 that has latest estimates for pre-pandemic year 2019, both with and without (w/o) or no shocks – unpredictable fatal discontinuities (war, famine, act of terrorism etc.) from Global Burden of Diseases, Injuries, and Risk Factors Study (GBD) 2019 *(6)*. The reported mortality rates for COVID-19 given in Table S1 are from (**8**).

The estimates of the mortality sex (Male: Female) ratio (MSR) for different countries as reported in **Table S2 & Table S3** were obtained by dividing mortality rates of males with that of females in RT-PCR confirmed cases, *i*.*e*., (mortality rate in males)/(mortality rate in females).

### METHODS

Basic data analysis including statistical calculations, *e*.*g*, Average, standard deviation (STDEV) presented as ±, 95% Confidence Interval (CI; shown as upper limit (UL) and lower limit (LL)) *etc*. performed in Excel 2019. Choropleth maps displaying cases sex ratio, mortality sex ratio (MSR), Fold Excess deaths (**Fig. 1-3**) were generated using Tableau Desktop 2021/2022.

#### Calculation: Table 1 and Tables S2 & S3

For calculation of quantities presented in Tables following formulae were used.

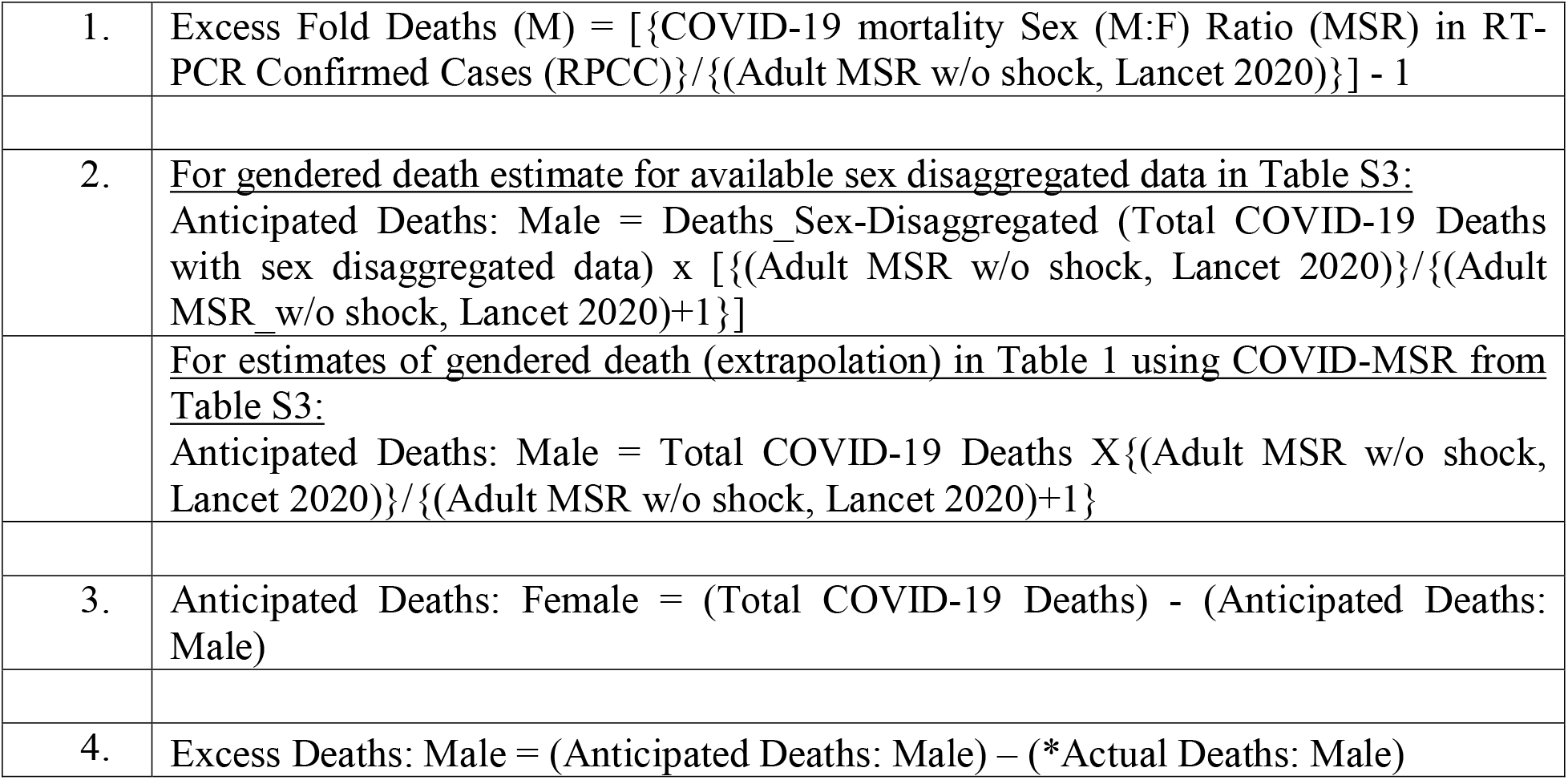
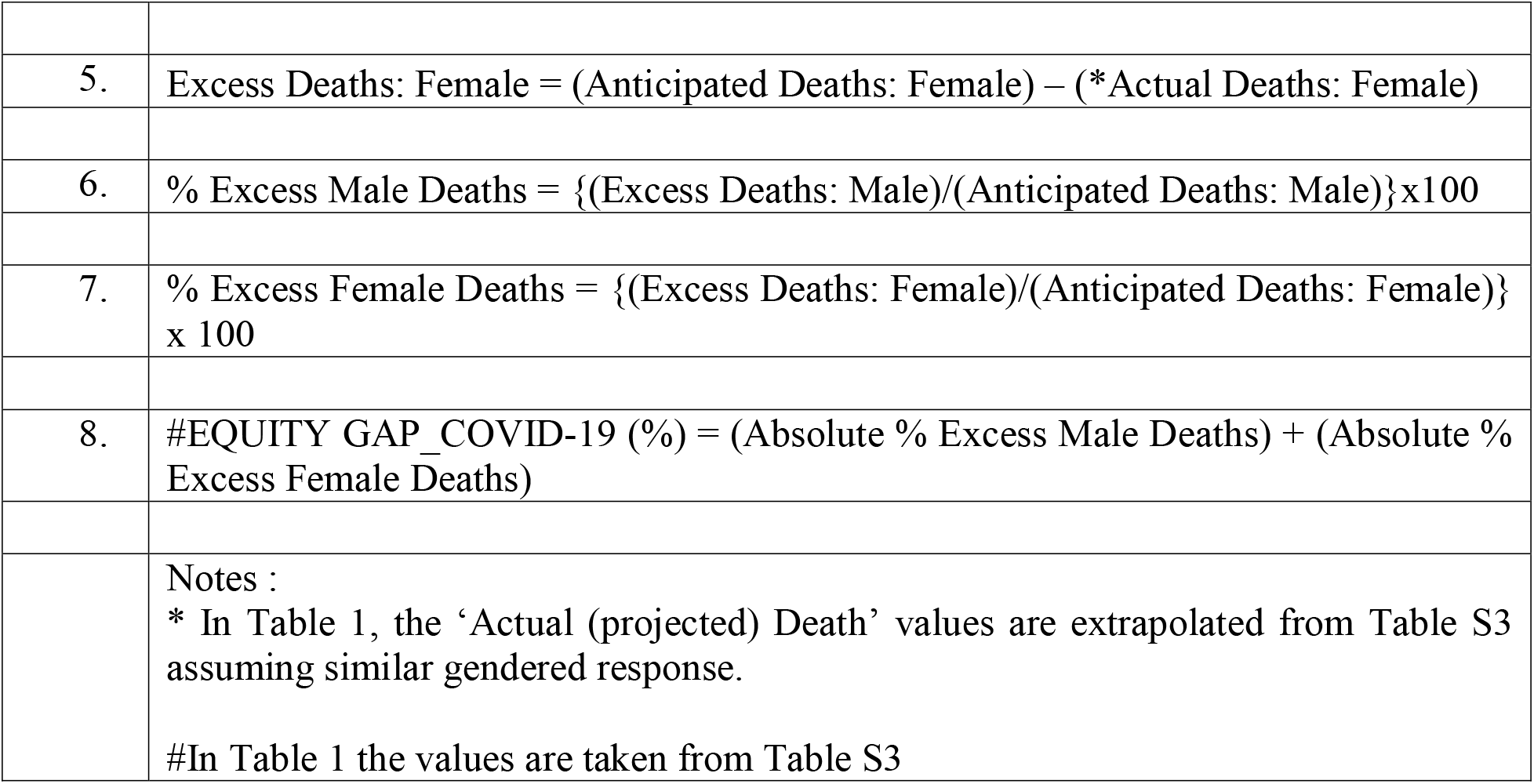

## Data Availability

All data produced in the present work are contained in the manuscript

## ACKNOWLEDGMENTS

The losses caused by the COVID-19 pandemic had been the source of inspiration for the work.

## AUTHOR CONTRIBUTIONS

Conceptualization: SS; Methodology & Investigation: SS, RKS; Visualization: SS; Writing – review & editing: SS, RKS

## DATA AND MATERIALS AVAILABILITY

All data are available in the main text, the supplementary materials, and sources”

## Supplementary Materials

Materials and Methods

Supplementary Text

Figs. S1 to S2

Tables S1 to S4

References *(23-27)*

