## Supplementary File for "Estimating the gendered impact of COVID-19"

**Includes Following**

Supplementary Methods & Text

Figs. S1 to S2

Tables S1 to S4

SUPPLEMENTARY METHODS & TEXT

***Gendered impact covariation with potential explanatory variables*:** The covariation of gendered deaths with some indices and variables previously modeled for explaining differential COVID -19 impact on populations are presented as **Figure S1 and Fig S2 (see supplementary text below).**

The indicators/indices and their sources used for the simple covariation analysis of the excess COVID-19 male/female mortality in countries were as follows:

(i) The data about Zn deficiency of populations is from Wessells and Brown, 2012 *(23)*.
(ii) The Latent Tuberculosis Infection (LTBI) prevalence estimate - measured as presence of reactivity against tuberculin (Tuberculin Sensitivity Test) a measure of exposure and presence of trained immunity against and as a result of *Mycobacterium tuberculosis* exposure (BCG vaccination or environmental isolates *Mycobacterium tuberculosis* complex) in the absence of active tuberculosis disease, is from Global Burden of disease study 2017*(24)*. LTBI is named as such for programmatic TB management purpose to indicate the existing risk of a small fraction of LTBI positive population to develop TB during their life time.
(iii) Healthcare Access and Quality (HAQ) Index is an indicator on national levels of personal healthcare access and quality based on 32 causes considered amenable to healthcare over time is based on Global Burden of Disease Study 2015 (GBD 2015) (IHME, 2017) *(25)*.
(iv) From *Our world in data* *(26)* (<https://covid.ourworldindata.org/data/owid-covid-data.csv>; Accessed 26 October 2021 https://github.com/owid/covid-19-data/tree/master/public/data/) the indicators obtained were namely, HDI: human_development_index (“A composite index measuring average achievement in three basic dimensions of human development—a long and healthy life, knowledge and a decent standard of living. Values for 2019, imported from http://hdr.undp.org/en/indicators/137506”), Life_Exp.@Birth: Life_Expectancy (Life expectancy at birth in 2019), Diabetes_Prev: Diabetes_Prevalence (Diabetes prevalence (% of population aged 20 to 79) in 2017), CVD_DR:cardiovasc_death_rate (“Death rate from cardiovascular disease in 2017 (annual number of deaths per 100,000 people”), Ext.Pov: Extreme_Poverty (“Share of the population living in extreme poverty, most recent year available since 2010”), Str.Ind.:Stringency_index [“Government Response Stringency Index: composite measure based on 9 response indicators including school closures, workplace closures, and travel bans, rescaled to a value from 0 to 100 (100 = strictest response)”]Pop. Density: Population_Density (“Number of people divided by land area, measured in square kilometers, most recent year available”), DpM: Deaths/Million (COVID-19 deaths/million population), CpM: Cases/Million (COVID-19 cases/million population) as on 26 October 2021, GDP per capita or GDP/capita (“Gross domestic product at purchasing power parity (constant 2011 international dollars), most recent year available”), Population (Population (latest available values). %Aged>65y (“Share of the population that is 65 years and older, most recent year available”), %Aged>70y (Share of the population that is 70 years and older in 2015). Additionally, the site https://github.com/owid/covid-19-data/blob/master/scripts/input/un/population_latest.csv may be assessed for full list of sources for different indices/values)
(v) Universal Health Coverage (UHC) Effective Coverage Index 2019 (Lancet 2020) and UHC TB Treatment coverage (UHC 2019_TB Treat. (Lancet 2020) based on Global Burden of Disease study 2019 (GBD 2019) *(27)*

***Supplementary Text***

Gendered Deaths display some interesting covariation with some of the potential population parameters presented (**Fig. S1 and S2**). The countries with > USD 14601 GDP/per capita, > 13% of population >65 years old (**Fig. S1**), and >0.8 HDI had exclusively displayed excess female deaths (**Fig. S2**) while it generally increased with increasing life expectancy for countries (**Fig. S2**).

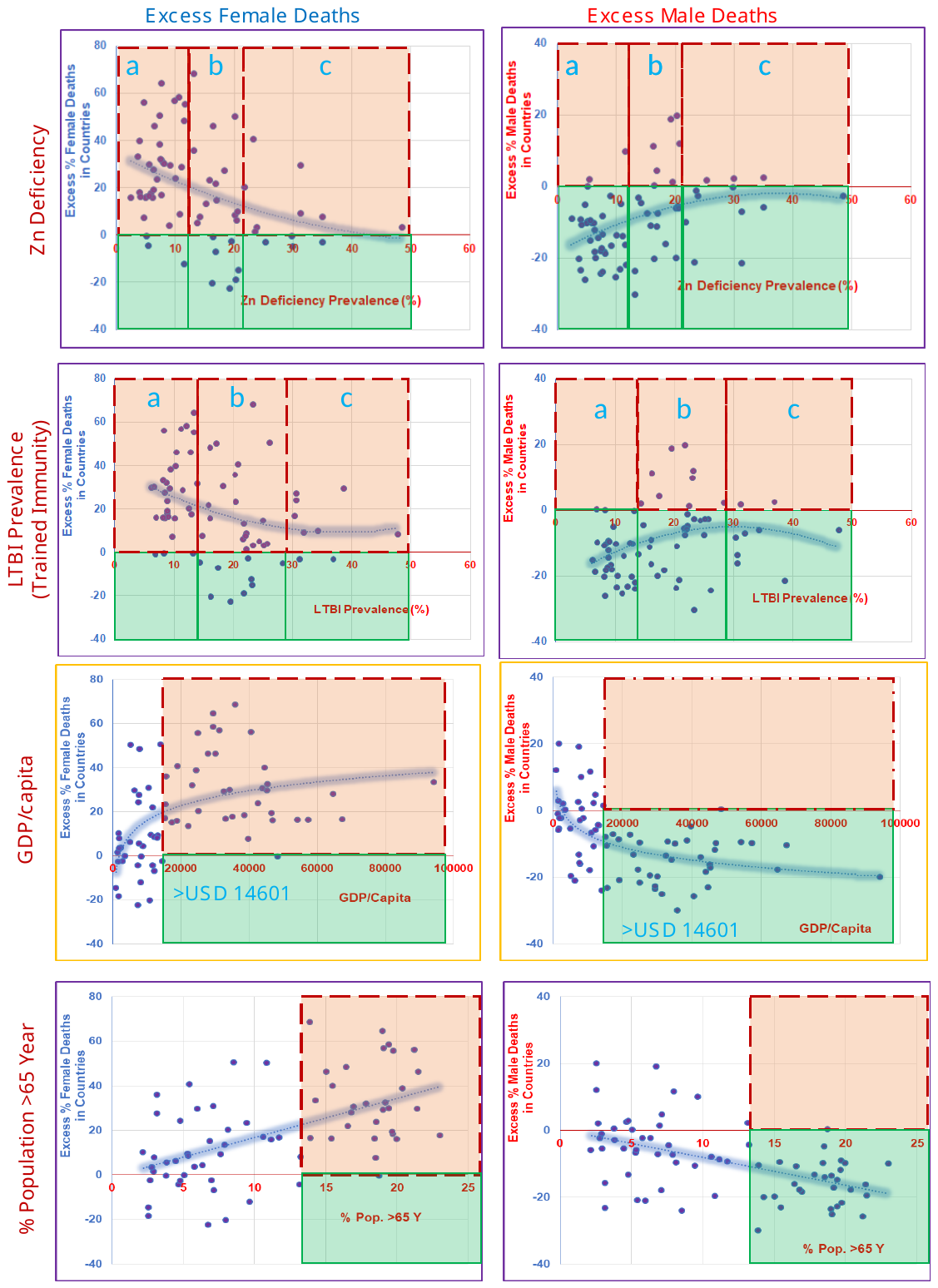

**Fig. S1 Gendered COVID-19 deaths – covariation with population parameters.** Note differential gendered deaths with variation in Zinc deficiency prevalence, LTBI prevalence, GDP/Capita and elderly and aged (>65 year) Population composition. Boxes bound by red dotted and green lines indicate countries with excess and reduced mortality, respectively.

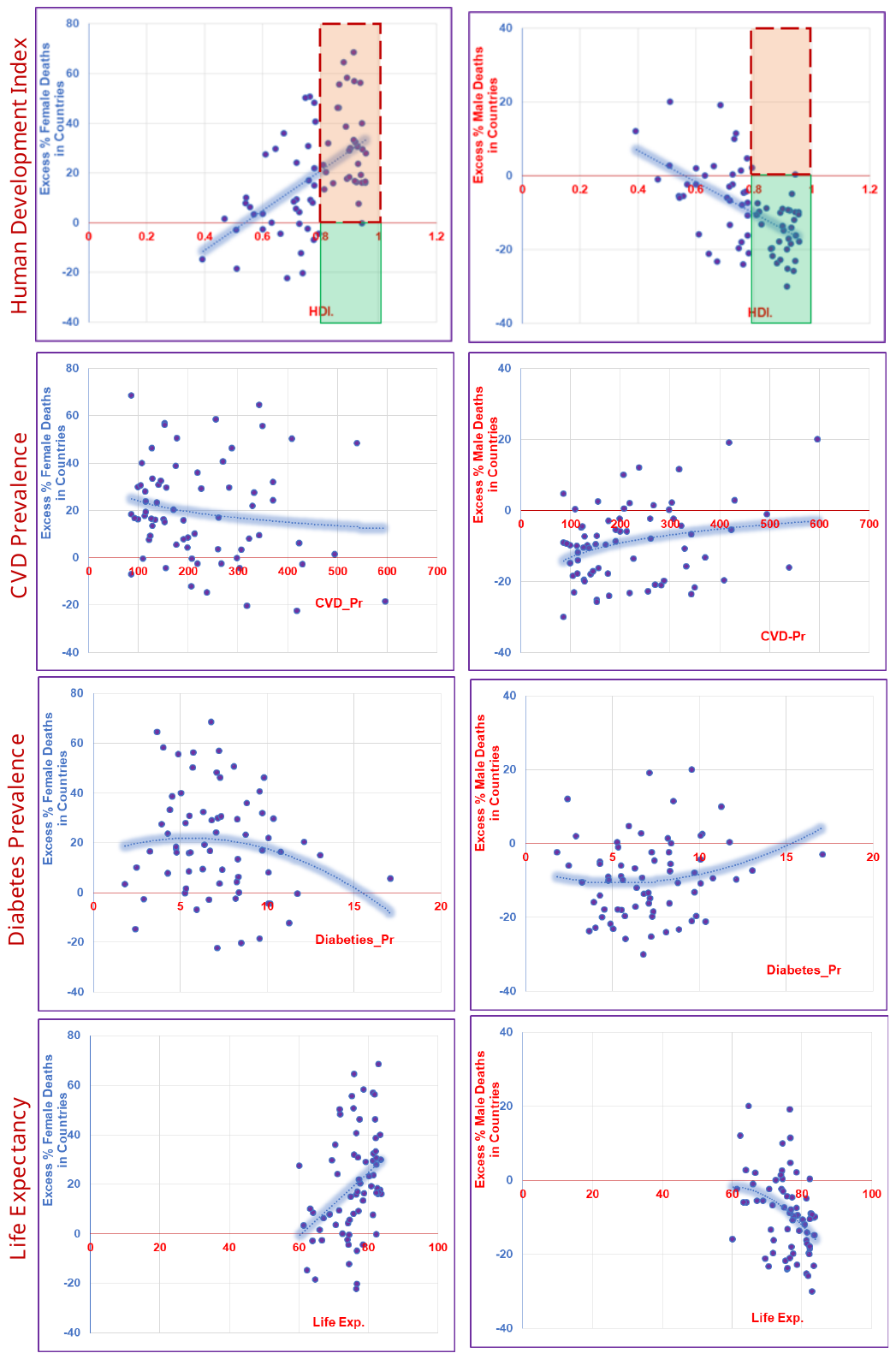

**Fig. S2. Gendered COVID-19 deaths - covariation with miscellaneous population parameters.** Note differential gendered deaths with Human Developmental Index, Cardiovascular Disease (CVD) prevalence, Diabetes prevalence, Life Expectancy. Boxes bound by red dotted and green lines indicate countries with excess and reduced mortality, respectively.

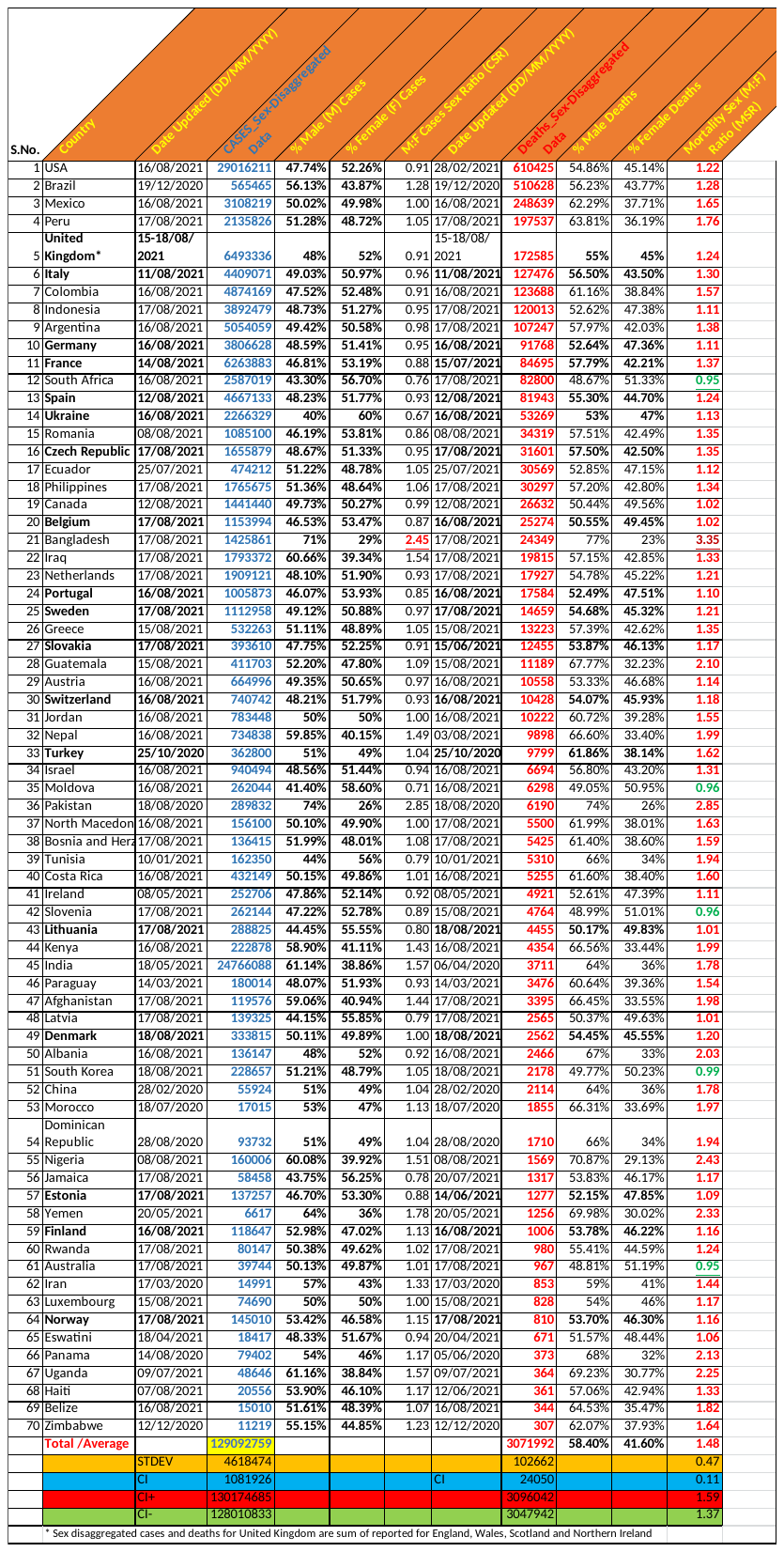

Table S1. The sex-disaggregated data of the top 70 countries providing data for at least >300 total COVID-19 deaths from Global Health 50/50 COVID-19 *(8)*. There seems to be excess female cases (*i.e.*, male(M): female(F) case sex ratio (CSR) <1.0 while excess male mortality (*i.e.*, male (M): female (F) mortality sex ratio (MSR) >1.0. The highest and lowest CSR and MSR are underlined. The date of submission/updating is indicated. Note the sex-disaggregated data submission had not been uniform across countries.

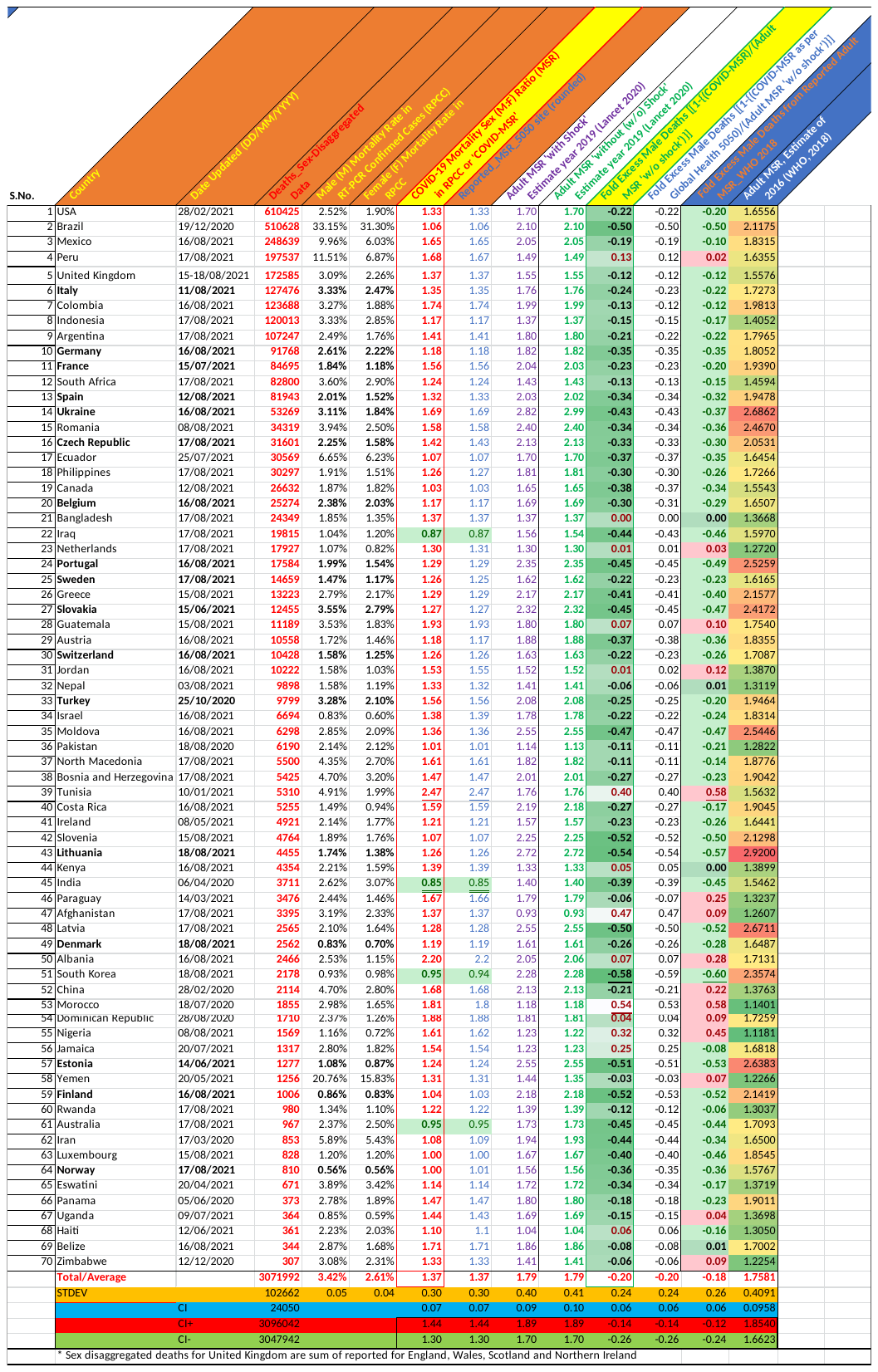

**Table S2.** C**ovid-19 mortality rates in RT-PCR confirmed COVID-19 cases were lower for males (higher for females) when adjusted with the prevailing mortality sex ratio (MSR) for adults.** Consideration of diagnostic gold standard the RT-PCR confirmed COVID-19 cases, indicates COVID-19 specific MSR lower than general adult MSR whether with or without discontinuities/shock. The estimated excess fold deaths of males, who are supposedly more disproportionately affected by COVID-19, come negative for the majority (56/70) of countries. In the majority of countries, the female mortality rate had been higher than expected**.**

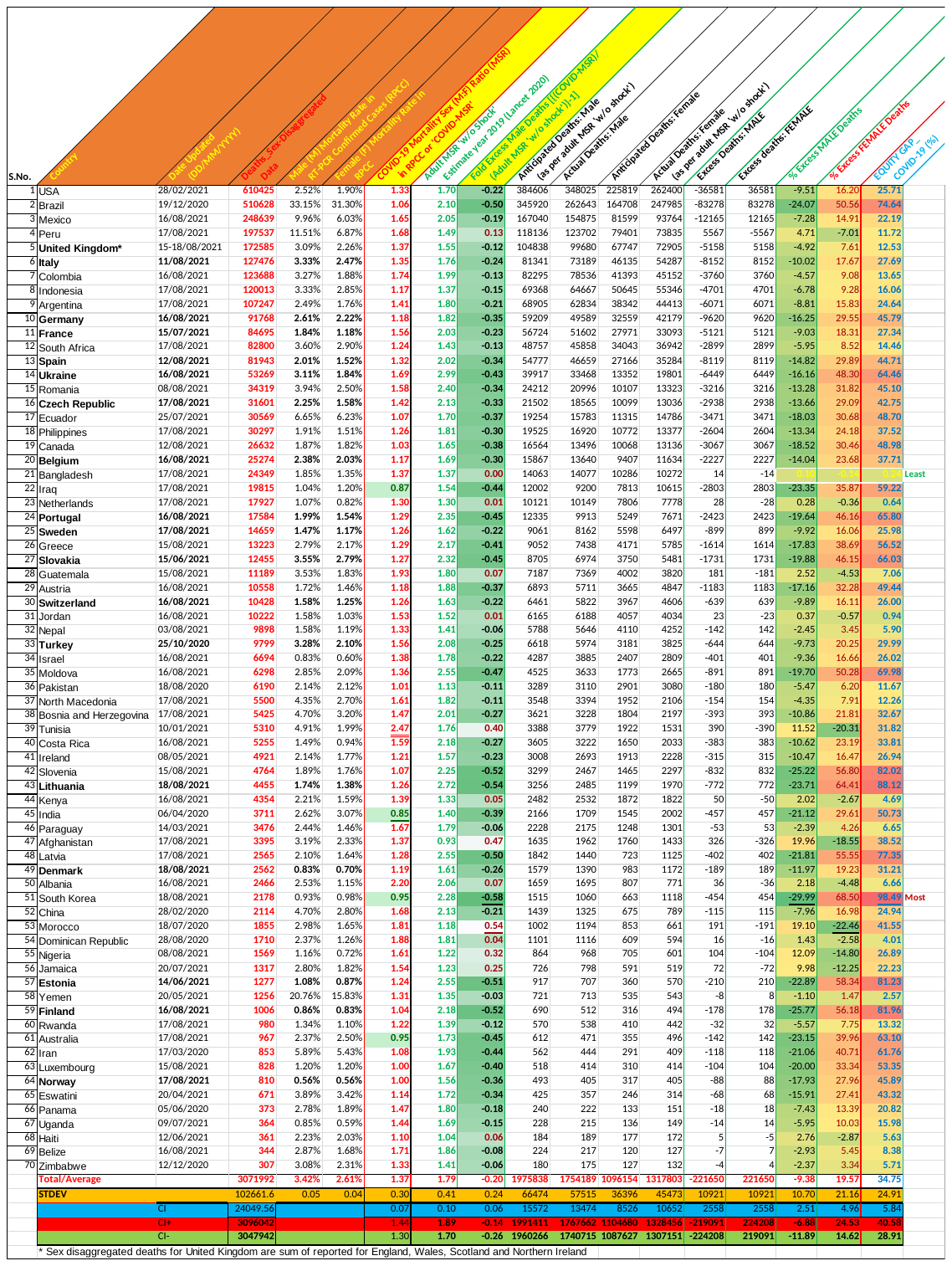

**Table S3. COVID-19 specific gendered mortality (anticipated and actual) and gender equity gap in COVID-19 response.** Male mortality (deaths) rates had been lower while female mortality (deaths) rates had been higher than would be anticipated from the country's prevailing pre-pandemic MSRs. Based on different MSR (w/o shock) for countries, the changed COVID-19 specific mortality sex ratio translates into a differential equity gap for countries. Surprisingly, Bangladesh and Netherlands appeared to be most gender-neutral in COVID-19 while South Korea followed by Lithuania, Slovenia, Finland, and Estonia displayed the largest equity gaps.

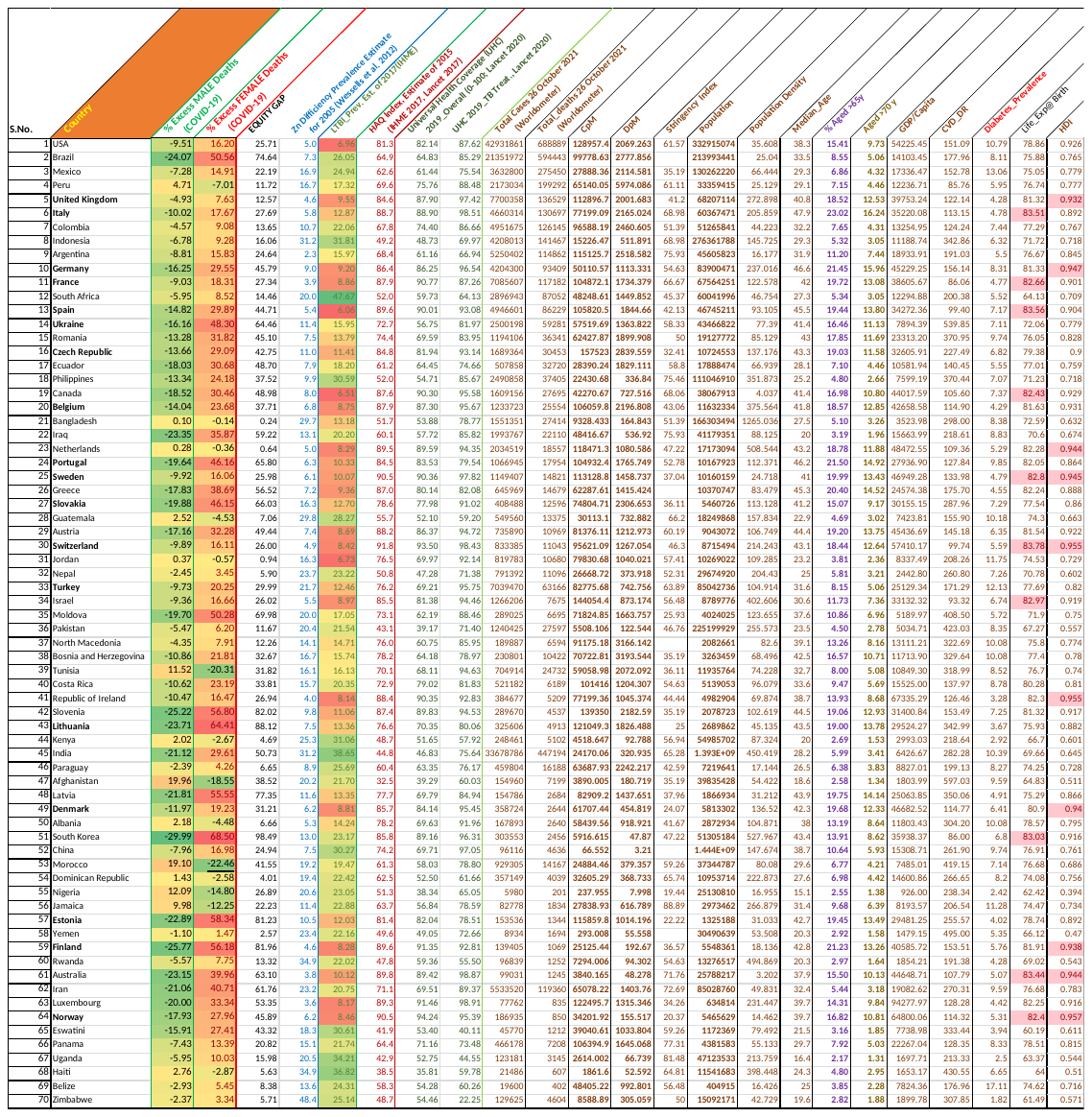

**Table S4. Gendered COVID-19 impact and potential variables**
